# Functional connectivity correlates of post-acute covid 19 cognitive and neuropsychiatric complications

**DOI:** 10.64898/2026.09.24.26363490

**Authors:** AM Auwal, S Keller, Ed Bullmore, MA Ellul, BD Michael

**Affiliations:** The WHO Collaborating Centre for Brain Health, Liverpool Interdisciplinary Neuroscience Centre, University of Liverpool, Liverpool, UK; The Brain Infection & Inflammation Group, University of Liverpool, Liverpool, UK; The Walton Centre NHS Foundation Trust, Liverpool, UK; King’s College London

**Keywords:** Regional Homogeneity, COVID-19, functional MRI, neuropsychiatry, biomarkers

## Abstract

**Background:** The neuropsychiatric sequelae (cognitive deficits, anxiety and depression) post-COVID-19 exist as part of the spectrum of the multisystemic long COVID. People with these symptoms have been shown to have worse functional outcomes. Hence, this significantly impacts the economy, primarily due to productivity losses. Yet, the precise underlying pathogenesis remains unknown, as do any diagnostic or prognostic biomarkers.

**Aim:** This study aims to identify functionally impacted brain regions in those with post-COVID-19 neurological sequelae and to evaluate how alterations in brain function relate to acute serum inflammatory mediators and brain injury biomarkers.

**Methods:** The study will utilise data from the COVID-19 Clinical Neuroscience Study (CNS), a large UK multicentre study conducted during the pandemic. This sub-study will be a secondary analysis of the COVID-19 CNS, a prospective observational study that compared COVID-19-positive participants with negative controls. Participants had an MRI, blood tested for acute serum inflammatory mediators and brain injury biomarkers, and neuropsychiatric assessment (cognitron test, PHQ-9 and GAD-7) post-acutely and at follow-up. The DPABI (Data Processing & Analysis for Brain Imaging) toolbox will be used for pre-processing and analysis of the neuroimaging data (resting state functional MRI) to obtain regional homogeneity (ReHo). The association between ReHO and neuropsychiatric symptoms, acute inflammatory mediators and brain injury biomarkers will be explored. Regression models would be used to determine the risk and protective factors for developing these symptoms.

**Conclusion:** It is anticipated that the study will provide new insights into the functional basis of post-acute COVID-19 neuropsychiatric manifestations and identify new diagnostic biomarkers.

## Background

Anxiety, depression and cognitive deficits were prevalent during the COVID-19 pandemic.^1–4^ Many years later, these symptoms persist in approximately 1 out of every 8 of those who had the infection, according to a prospective cohort study in the Netherlands that followed 1,782 COVID-19-positive and 4,130 controls.^5^ It was later reported that this cohort with cognitive and neuropsychiatric complications had worse functional outcomes related to impairment in activities of daily living and impact on employment, resulting in^6^ a global annual economic burden of $ 1 trillion, mainly attributable to productivity losses.^6 5^Although these neuropsychological and neuropsychiatric features were thought to be due to the neurological consequences of COVID, they could also be explained or exacerbated by sociocultural factors, such as social isolation, lifestyle changes and deteriorating living conditions associated with the pandemic, suggesting the need to investigate their biological basis.^7,8^

Several theories have been put forward to explain the pathogenesis of these cognitive and neuropsychiatric symptoms post-COVID-19, including neuroinflammation via immune dysregulation and autoimmunity, neuronal injury, dysfunctional neurological signalling, endothelial dysfunction and coagulopathy, gut microbiome dysbiosis, but none have been conclusive.^9–11^ Brain injury biomarkers (Nfl, GFAP, UCHL1, tau) and inflammatory mediators (IL-6, IL-1RA, IL-12p40, HGF, M-CSF and CCL2) have previously been reported to be elevated in those with neurological complications of COVID-19, but whether they have any significance on functional connectivity in this group is yet to be explored.^12^ Although the pathogenesis of post-COVID-19 cognitive and neuropsychiatric manifestations remains unclear, certain factors have been associated with an increased risk of developing these complications, including female sex, being over 50 years, BMI > 25 kg/m^2^, type 2 diabetes, multimorbidity, hospitalisation or ICU admission related to COVID-19. In contrast, vaccination and treatment with dexamethasone were protective.^13–16^

In addition to molecular studies, several structural and functional neuroimaging studies have attempted to elucidate the neuroimaging correlates of post-COVID-19 cognitive and neuropsychiatric sequelae and examine whether they will explain the observed cognitive and psychiatric deficits.^14,17,18^ The UK Biobank study, for example, identified a reduction in grey matter thickness in the parahippocampal gyrus, orbitofrontal cortex, and global brain volume in COVID-19 positive participants compared to healthy COVID-negative controls.^17^ Greta and colleagues demonstrated a reduction in anterior cingulate cortex volume which correlated with cognitive deficits across multiple domains in COVID-19-positive participants compared to matched COVID-19-negative controls.^14 18^

Functional connectivity, which is the temporal correlation between spatially distinct brain regions, is useful in elaborating inter-regional associations but often relies on a priori selection of regions of interest, meaning it is susceptible to missing other significant regions.^19^ Regional Homogeneity (ReHo) on the other hand measures how an individual voxel relates to its neighbours over time. It explores all brain regions and can detect subtle local changes, hence, a sensitive way of studying the influence of other factors on brain connectivity.^20^ It is, however, also influenced by cerebral blood flow and cardiovascular risk factors. Several studies have shown functional alterations in COVID-19-positive participants compared to COVID-19-negative controls and how these alterations correlated with cognitive and neuropsychiatric complications.^18^ These are, however, far from being used as biomarkers due to the varied regions these connectivity changes involve and non-uniformity in their directionality. Thus, combining ReHo and functional connectivity will be a robust means to study functional alterations in post-COVID-19-related cognitive and psychiatric complications.

A search in publication databases revealed very few studies which looked at local connectivity in COVID-19 out of which only two studied ReHo in COVID-19.^21–23^ Of these two, one of the studies demonstrated decreased ReHO in the left superior temporal gyrus within COVID-19 survivors two years after compared to healthy controls, and that this decrease negatively correlated with cognitive function assessed by the abbreviated Cognitive Failures Questionnaire-14.^23^ The other study showed reduced ReHo within COVID-positive patients when compared to healthy controls in the anatomically close left angular gyrus, and this negatively correlated with depressive symptoms assessed using the Patient Health Questionnaire (PHQ-9). Higher ReHo was, however, seen in the right hippocampus, which positively correlated with the PHQ-9 scores.^21^ Although these studies showed ReHO alterations in closely related brain regions, they correlated with different clinical outcomes. This is probably related to the differences in sample size and timing of assessment post COVID-19 between the two studies. Our study will be more robust with a larger cohort, combining ReHO with seed-based functional connectivity, to give a clearer insight into the nature of functional changes in post-acute COVID-19 patients and how these relate to cognitive and neuropsychiatric sequelae. It will additionally determine how these ReHo changes relate to acute inflammatory and brain injury biomarkers.

### Study Proposal

In this multicentred UK National study, we aim to identify the functional correlates of cognitive and neuropsychiatric complications of post-acute COVID-19 to better understand their pathophysiology and explore biomarkers that could be prognostic or serve as targets for therapeutic intervention.

### Aim

To identify the functional correlates of cognitive and neuropsychiatric complications of post-acute COVID-19 patients using ReHo and seed-based functional connectivity, and whether these are associated with raised serum inflammatory and brain injury biomarkers.

### Hypotheses

1. Patients with neurological complications of COVID-19 (NeuroCovid-19) will show functional alterations post-acutely in the orbitofrontal cortex, insula, cingulate cortex, hippocampus, superior/middle temporal gyri and angular gyrus when compared to COVID-19 patients with no neurological complications or COVID-19 negative controls
2. The observed functional alterations post-acutely in NeuroCOVID-19 patients will correlate with cognitive dysfunction, anxiety and depression
3. Functional alterations post-acutely in NeuroCOVID-19 patients will correlate with elevated serum inflammatory and brain injury markers

### Outcomes

#### Primary

Determine the functional correlates of anxiety, depression and cognitive dysfunction post-acutely in NeuroCOVID-19 patients

#### Secondary

Identify potential inflammatory and brain injury biomarkers that are associated with functional correlates of cognitive and neuropsychiatric complications post-acutely in NeuroCOVID-19 patients

## Methodology

### Study design

This is a secondary analysis of a prospective cohort study done between post-acute COVID-19 patients and COVID-19 negative controls.

### Comparison groups

Post-acute COVID-19 with neurological complications (NeuroCOVID)

Post-acute COVID-19 without neurological complications

Negative COVID-19 controls

### Eligibility criteria

#### Inclusion

Adults > 16years of age

Meeting the WHO case definition of definite or probable COVID-19^24^ Having had a functional magnetic resonance imaging (fMRI)

#### Exclusion

Pre-existing psychiatric and/or neurological disorder Neurological complications from the COVID-19 vaccination Taking psychotropic medications at the time of the study

### Participants

Participants were recruited between March 2021 and October 2022 as part of the COVID-19 CNS study across 17 UK sites within the National Institute for Health Research (NIHR) COVID-19 BioResource (***Figure 1*** below). Healthy controls who had neuroimaging before the pandemic were recruited from the NIHR Bioresource. Those who met the eligibility criteria as detailed above will be included in this sub-study.

**Figure 1:**
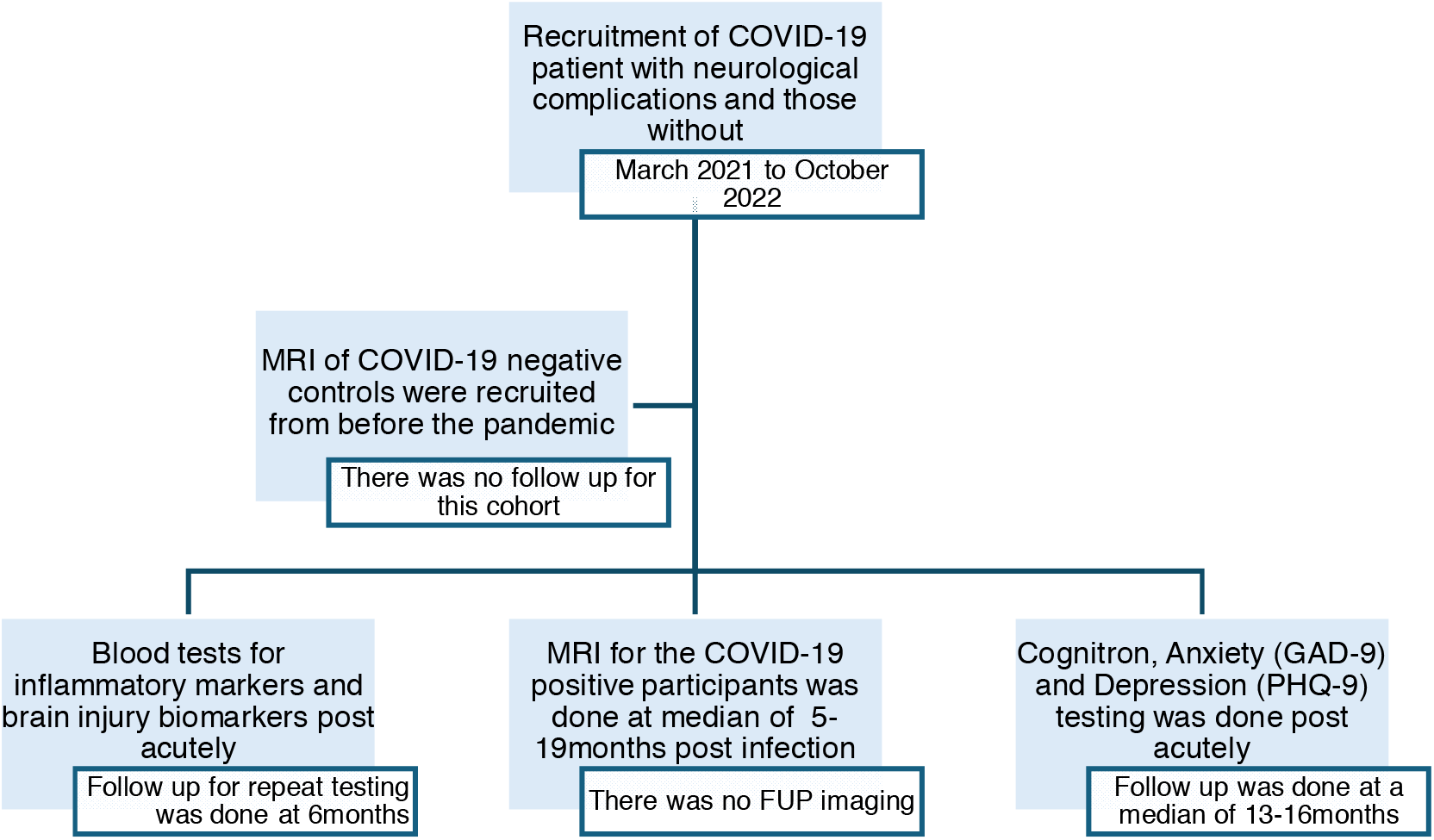
Flowchart for the timeline of recruitment into the COVID-CNS Study.

### Sample size justification

This utilises an unpaired t-test in G Power v 3.1

#### Based on Cognition

Hampshire and colleagues demonstrated that COVID-19 survivors were less accurate (G_SScore=-0.53SDs) than matched controls on the cognition test. Based on this, 58 participants in each group are sufficient to provide a study power of 80% in detecting a difference with one-tailed alpha at < 0.05.

#### Based on ReHo

Z-scores derived from ReHo at the hippocampus from *Catterineusi 2022* were –0.580 and – 1.020 for the COVID-19 and healthy groups, respectively. This gives Cohen’s d of 0.44, translating to 65 participants in each group at 80% power and 0.05% significance level. ^21^

#### Based on inflammatory markers

Michael and colleagues demonstrated statistically significantly higher levels of Nfl in COVID-19 patients with neurological complications (62.92 ± 179.19) compared to COVID-19 patients without neurological complications (22.43 ± 13.25) or COVID-19 negative controls (17.22 ± 15.72). ^25^ Stemming from this, 121 patients from each group will be required to have 80% power at the 0.05% significance level.

### Neuroimaging

#### Imaging acquisition

MRI was performed post-acutely across 4 different UK sites using 3 Siemens PRISMA scanners (Liverpool, Oxford, Cambridge) and 1 GE scanner (King’s College London).

Multiple sequences including T1 weighted (magnetisation prepared rapid gradient echo using Siemens; acquisition time = 4:54; resolution = 1.0×1.0×1.0; matrix = 256×256×208; TI/TR = 800/2000 ms; R = 2; this is similar for GE except that the acquisition time here is 4:42) and resting state functional MRI (acquisition time = 7:00; resolution = 2.4×2.4×2.4; matrix = 88×88×64, TE/TR = 39/735 ms, flip angle = 52°, MB = 8; for both Siemens and GE except that the acquisition time in the latter = 7:21) were obtained. The reliability of these multi-modal and multicentred MRIs has been validated in a previously published ‘travelling heads study’.^26^

#### Image pre-processing

The preprocessing of structural and functional magnetic resonance imaging data will be done using Data Processing and Analysis for Brain Imaging toolbox (https://rfmri.org/DPABI) and statistical parametric mapping (SPM12 or later) (https://www.fil.ion.ucl.ac.uk/spm/software/) on MATLAB (The MathWorks Inc., Natick, MA, US) interface (R2024b or later). Images will be inspected for structural anomalies at the beginning. Volume slices will be adjusted to account for the varying acquisition times. This will be followed by realigning the time series of images for each subject. Head motion will be corrected using the Friston 24-parameter model (Friston et al. 1996). The resulting structural images will be partitioned into Grey Matter (GM), White Matter (WM) and Cerebrospinal Fluid (CSF). The GM will then be transformed to the Montreal Neurological Institute (MNI) Space. ^27^

##### Regional homogeneity analysis

DPABI will be used for ReHO analysis. A cluster will be formed by any given voxel and its nearest 26 neighbours such that the time series of that voxel and its neighbours will be defined by Kendall’s coefficient of concordance (KCC). An individual ReHO map will be generated and then divided by the participant’s global mean KCC value within the brain mask for standardisation. The ReHO maps will then be partially smoothed (full width at half maximum = ?6mm) in SPM12. A two-sided sample t-test will be conducted to compare smoothed ReHO maps.

Removing the influence, alteration in brain volume might have on ReHO will be done on 2 levels. Firstly, the eigenvariate of the Grey Matter Volume (GMV) for each significant cluster will be estimated across segmented, normalised and modulated GM images in each study participant. Secondly, total intracranial volume (GM+WM+CSF volumes) and total GMV will be compared to exclude the global effects of brain volumetry. Differences in ReHo between NeuroCOVID, COVID and negative controls will be observed in those with elevated serum inflammatory and brain injury markers and those without.^27^

##### Neuropsychiatric assessments

Participants completed validated cognitive and psychological assessments post-acutely and at follow-up.

The cognitive test was based on the validated Cognitive Assessment Battery (Cognitron), which assesses five domains: complex attention, executive function, perceptual-motor control, language, and learning and memory. Depression was assessed using the validated Patient Health Questionnaire-9 (PHQ-9) and anxiety with Generalised Anxiety Disorder–7 (GAD-7). ^28–34^

### Serum and brain injury markers measurement

Blood was collected from both the participants and controls post-acutely and at follow-up. This was assayed for inflammatory mediators using Luminex and brain injury markers using Quanterix Simoa device.

### Statistical analyses

The analysis will be done in R version 4.5.0 or later.

The data distribution will be visually tested using Q-Q plots and the Kolmogorov-Smirnov test to direct the choice of an appropriate statistical test when comparing groups.

Homogeneity of variance will be tested using Bartlett’s test for normally distributed data and the Levene’s test for data that is not normally distributed to avoid underestimating the level of significance, which can lead to a type 1 error when overestimated or a decrease in power when underestimated.

FDR will be used to adjust for the level of significance when multiple comparisons are being done to reduce the risk of false positives, which may inflate type 1 error.

#### Analysis 1

Hypothesis:

1 Covid-19 patients would show more ReHo alterations in areas associated with anxiety, depression, fatigue and cognitive impairment than healthy controls
- An independent T-test in SPM will be used to compare Kendall’s Coefficient of Concordance (KCC) between COVID-19 cases and controls, which gives a measure of ReHO
- Total volumes (GMV + TIV) with age as covariates or local volumes with age and TIV (Total Intracranial Volume) as covariates will be compared using ANCOVA to evaluate for any influence that change in brain volume might have on ReHO.

#### Analysis 2

2 ReHo alterations in COVID-19 patients will correlate with neuropsychiatric symptoms
3 Changes in ReHo in post-acute COVID-19 patients are associated with raised acute inflammatory and brain injury biomarkers
- Pearson’s correlation will be used if the data is normally distributed, or Spearman’s rank correlation if otherwise, to determine any association between the inflammatory/brain injury biomarker and the observed ReHO. This will give insight into the mechanistic process of the observed neuropsychiatric manifestations in post-acute COVID-19 and potentially identify a diagnostic biomarker.

#### Analysis 3

4 Time since COVID, severity, and multimorbidity are risk factors for developing neuropsychiatric symptoms, whereas vaccination and prior treatment with dexamethasone may be protective.
- A multiple regression model for each neuropsychiatric symptom will be developed to identify both risk and protective factors. This is preferable to tests such as simple regression, in that it is more precise in examining a complex relationship between multiple predictor variables while controlling for confounding variables.
- Independent variables will be: Age, time since COVID, severity, multimorbidity, educational level, vaccination status and treatment with dexamethasone
- Correlation between independent variables will be tested, and those found to be highly correlated will be removed

## Results

This will be reported in accordance with the “Strengthening the Reporting of Observational Studies in Epidemiology (STROBE)” framework.

### Impact

The study is set to determine the neural correlates of the neuropsychiatric manifestation in post-acute COVID-19 patients, address the mechanistic gap and identify diagnostic/prognostic biomarkers.

### Conclusion

This study aims to identify the functional neural correlates of the neuro-psychiatric consequences post-COVID-19 and whether these are associated with acute serum inflammatory or brain injury biomarkers. It will also give insight into the risks and protective factors against developing these sequelae. This will consequently help address the mechanistic gap in long COVID and identify diagnostic/prognostic biomarkers.

### Strengths

The study has high power and is enriched with various data (blood workup, neuroimaging, and neuropsychiatric testing). There is no a priori selection of regions for the intended analysis of the functional MRI using regional homogeneity.

### Limitations

This includes the lack of standardisation of the time at which the various data forms were collected. Additionally, regional homogeneity can be very susceptible to noise from physiological functions such as respiration and cardiac activity, which may lead to the misinterpretation of the findings.

## Data Availability

Not applicable since this is a protocol

## Ethics

Next of kin or participants, where possible, provided consent. The study was registered with the East of England Cambridge research ethics committee with reference number: REC reference 17/EE/0025; 22/EE/0230

